# Blood Pressure Variability in Daily Life: Comparing Individuals with Cervical and Upper-Thoracic Spinal Cord Injury to Non-Injured Controls

**DOI:** 10.64898/2025.12.17.25342472

**Authors:** Shane J.T. Balthazaar, Matthias Walter, Catherine R. Jutzeler, Andrea L. Maharaj, Andrei V. Krassioukov, Tom E. Nightingale

**Affiliations:** School of Sport, Exercise and Rehabilitation Sciences, College of Life and Environmental Sciences, University of Birmingham, Edgbaston, Birmingham, United Kingdom (UK); Institute of Cardiovascular Sciences, University of Birmingham, Edgbaston, Birmingham, UK; International Collaboration On Repair Discoveries (ICORD), Faculty of Medicine, University of British Columbia, Vancouver, British Columbia (BC), Canada; Department of Cardiology, University Hospitals Birmingham National Health Service (NHS) Foundation Trust, Birmingham, UK; Faculty of Medicine, University of Basel, Basel, Switzerland; Department of Health Sciences and Technology, ETH Zürich, Zürich, Switzerland; SIB Swiss Institute of Bioinformatics, Lausanne, Switzerland; Division of Physical Medicine and Rehabilitation, Faculty of Medicine, University of British Columbia, Vancouver, BC, Canada; GF Strong Rehabilitation Centre, Vancouver Coastal Health, Vancouver, BC, Canada

**Author notes:** Tom E. Nightingale, School of Sport, Exercise and Rehabilitation Sciences College of Life and Environmental Sciences, Y14 - University of Birmingham, Edgbaston, Birmingham, UK, B15 2TT, Andrei V. Krassioukov, ICORD-BSCC, UBC, 818 West 10th Avenue Vancouver, BC, Canada, V5Z 1M9.

**Keywords:** cardiovascular diseases, hypotension, hypertension, circadian rhythm, ambulatory

## Abstract

**Background:** Spinal cord injury (SCI) above the sixth thoracic spinal cord segment can disrupt autonomic cardiovascular control, leading to increased BP variability (BPV). This study compared BPV, nocturnal dipping patterns, and blood pressure (BP) fluctuations among individuals with cervical SCI (C-SCI), upper-thoracic SCI (UT-SCI), and non-injured controls (NI-C).

**Methods:** Using 24-hour ambulatory blood pressure monitoring, we analyzed BPV (via standard deviation [SD], coefficient of variation [CoV], average real variability [ARV], and variability independent of the mean [VIM]), nocturnal dipping, and systolic/diastolic BP fluctuations. Nocturnal dipping percentages were calculated and patterns classified. Hypotensive (SBP <100mmHg, DBP <70mmHg) and hypertensive (SBP >150mmHg per clinical guideline thresholds) events were identified. BP distribution was analyzed using skewness and kurtosis.

**Results:** Eighty participants (44 C-SCI, 19 UT-SCI, 17 NI-C) had 66 ± 18 measurements taken and are included in the analysis. The C-SCI group exhibited significantly higher systolic BPV metrics across SD, CoV, ARV, and VIM compared to the UT-SCI and NI-C groups (*P* < 0.01). C-SCI nocturnal BP dips were reduced, and reverse dipping patterns were more prevalent (*P* < 0.001). Hypotensive events occurred more frequently in C-SCI and UT-SCI compared to NI-C (both *P* < 0.001). SCI groups, particularly those with C-SCI, showed significantly more right-skewed SBP distributions and a higher proportion of non-Gaussian BP profiles, suggesting increased BP lability.

**Conclusion:** Individuals with C-SCI showed significantly increased BPV and impaired nocturnal dipping, while both C-SCI and UT-SCI demonstrated heightened susceptibility to hypotensive events. These findings highlight the need for targeted cardiovascular monitoring and interventions in individuals with C-SCI and UT-SCI to mitigate the known risks associated with BP dysregulation.

## Introduction

Spinal cord injury (SCI) often leads to a complex array of impairments, encompassing both sensorimotor and autonomic dysfunction.^1–3^ Disruptions of autonomic cardiovascular control leads to blood pressure (BP) abnormalities,^4^ increasing cardiovascular disease (CVD) risk.^5^ Injuries above the sixth thoracic spinal cord segment (T6) present challenges due to the loss or disruption of descending sympathetic control over the heart, trunk and lower-extremity vasculature.^6^ This disruption may lead to complications such as autonomic dysreflexia (AD)^7^ and hypotensive episodes triggered by exercise,^8^ orthostatic challenges,^9^ or postprandial states.^10^ The heart receives dual innervation – parasympathetic input via the vagus nerve and sympathetic input from T1-T5 segments of the spinal cord.^11^ However, most blood vessels rely solely on sympathetic innervation, with the exception of blood vessels in the genitalia.^12^ This selective innervation of target organs and variability in the degree of descending sympathetic vasomotor control contributes to unique cardiovascular responses and complexities in BP regulation within the SCI population.^13^ Notably, some individuals with SCI remain asymptomatic during AD and hypotensive events, highlighting the importance of thorough assessment and management.^14^ From a pathophysiological perspective, greater BP variability (BPV) is expected in cervical SCI (C-SCI) due to loss of supraspinal sympathetic cardiac control compared with upper thoracic SCI (UT-SCI; T1-T6), where partial cardiac regulation may persist.

Blood pressure naturally fluctuates in response to environmental, physical, and emotional stimuli.^15^ While averaging multiple BP readings is a standard practice, this approach may not adequately capture the dynamic BP patterns observed in individuals with SCI, as autonomic dysregulation causes greater BPV, exaggerated responses to stimuli, and prolonged hypertensive or hypotensive episodes.^16–18^ Blood pressure variability, defined as a change in the value of arterial blood pressure over a defined period of time,^19^ has emerged as a critical parameter in cardiovascular research. Elevated BPV is associated with end-organ damage, stroke, and mortality, independent of mean BP.^20–22^

Ambulatory blood pressure monitoring (ABPM) provides valuable insights into BP lability in SCI,^23^ revealing patterns with implications for cardiovascular morbidity and mortality.^24^ Prior studies show disrupted circadian BP profiles,^25,26^ particularly among individuals with motor-complete tetraplegia.^24^ In healthy individuals, BP typically declines by 10-20% during sleep due to reduced sympathetic activity.^27^ Variations in nocturnal BP patterns include extreme dippers (BP drops >20%), dippers (BP drops 10-20%), non-dippers (BP drops <10%) and reverse dippers (BP increases at night).^28,29^ These classifications are clinically important, as reverse dipping is linked with increased CVD risk.^30^

Despite the potential of ambulatory BPV indices to capture differences in a free-living environment, their utility in a large SCI cohort compared with age-matched controls remains unclear. Frisbie and colleagues reported greater short-term BP instability in complete cervical SCI compare to low thoracic injuries, suggesting that higher injuries cause greater baseline BP volatility independent of posture or other triggers.^31^ Similarly, Munakata and colleagues demonstrated attenuated or absent nocturnal BP dipping and exaggerated BP fluctuations in motor-complete tetraplegia, indicating profound autonomic dysregulation even without overt cardiovascular symptoms.^32^ Rosado-Rivera and colleagues extended these findings by comparing 24-hour BP and autonomic profiles across individuals with tetraplegia, high and low-level paraplegia, and controls, showing lower 24-hour systolic BP and blunted nocturnal dipping in individuals with tetraplegia.^33^ Collectively, these studies confirm that SCI alters circadian BP regulation and increases BP lability but have not comprehensively quantified multiple BPV indices.

Therefore, this study expands on previous work by pooling data from four independent cohorts to enable stratification by neurological level of injury and comprehensive quantification of time-domain BPV metrics. While these indices (described in detail in the Methods) are well established in non-injured populations,^34^ they remain underexplored in individuals with SCI. By integrating these indices, this study aims to clarify how injury level influences BPV patterns and to establish a foundation for assessing their potential prognostic relevance for cardiovascular risk in this population.

## Methods

### Study Design and Ethical Considerations

This study is a secondary analysis of baseline data obtained from three longitudinal clinical trials (registration identifiers NCT02298660 (April 2013 – December 2017),^35^ NCT02676154 (April 2016 – January 2019),^36^ and NCT01718977 (April 2013 – April 2019),^37^ and a cross-sectional study, all approved by the Clinical Research Ethics Board (CREB) at the University of British Columbia. Detailed information describing the objective of each of these trials and the trial specific inclusion and exclusion criteria can be found in Supplementary Table S1. Participants with SCI were assessed using the 2011 version of the International Standards for Neurological Classification of Spinal Cord Injury (ISNCSCI), a standardized neurological examination used to determine neurological level of injury (NLI) and classify the severity of motor and sensory impairment.^38^ The severity of impairment was categorized according to the American Spinal Injury Association (ASIA) Impairment Scale (AIS), which ranges from AIS A (sensorimotor complete injury) to AIS D (sensorimotor incomplete injury). Inclusion criteria for all participants in this study: adults aged 18 to 60 years with a chronic (> 1-year), traumatic SCI at or above T6. Both males and females were eligible to participate, provided they were competent to give informed consent. Exclusion criteria for all participants in this pooled analysis included a history of cardiovascular or significant cardiopulmonary diseases, severe acute or unstable medical conditions, significant renal or hepatic dysfunction, recent major trauma or surgery within the past six months, or those with uncontrolled psychiatric or substance abuse issues. Additionally, women who were pregnant or breastfeeding were excluded. Age and sex-matched non-injured control (NI-C) data from the cross-sectional study were collected for comparison to the SCI participants.

### Ambulatory Surveillance Protocol

All enrolled participants underwent a 24-hour ambulatory surveillance period, encompassing continuous monitoring of brachial BP and HR (24-hour ABPM) using either the Meditech Card (X)plore device (Meditech Ltd., Budapest, Hungary) or the Mobil-O-Graph (IEM, Stolberg, Germany), which have both been clinically validated for use.^23,39^ These devices have published protocol validations confirming the accuracy of brachial BP measurements (i.e., the basis for calculating time-domain BPV).^40^ Although two device types were used across the pooled cohorts, all sites followed a uniform acquisition protocol and the same data analysis framework. Time-domain BPV indices are insensitive to fixed calibration differences, and standardizing sampling schedules and preprocessing steps helps minimize potential device-related effects.^41^ BP and HR were set to record at least every 15 minutes from 9:00AM until 8:00PM, then at least hourly between 8:00PM to 9:00AM, with adjustments based on participants’ self-reported sleep and wake times. Across all study protocols, participants (C-SCI, UT-SCI, and NI-C) were also encouraged to take additional BP and HR readings at any time if they experienced any symptoms of low or high BP or engaged in any activities that might influence BP (e.g., urinary bladder and bowel management, physical exertion, moving to an upright position, taking medications, eating, etc). To summarize each participant’s typical BP, we used the median 24-hour systolic BP (SBP) and diastolic BP (DBP). Ambulatory BP in chronic SCI is often skewed and characterized by episodic excursions. The median therefore provides a robust measure of central tendency that reflects a participant’s usual BP without being disproportionately shifted by brief, clinically meaningful spikes or drops in BP. These extreme values were not removed from analysis; they were incorporated into all BPV indices, which were computed from the full time series. As a sensitivity check, all analyses were repeated using the mean 24-hour SBP and DBP, with no material change in results. Considering the real-world challenges of monitoring ambulatory BP the SCI population,^24,42^ a valid measurement was defined as having at least 60% of attempted BP readings successfully recorded (i.e., an attempted BP measurement that gives an accurate reading and not an error warning) during the 24-hour monitoring period. This threshold ensured sufficient data to reliably represent each participant’s BP patterns and allow for meaningful analysis. To assess BPV, which has been used as a marker for CVD risk in prior studies, statistical measures such as the standard deviation (SD) of an individual’s BP readings can be used.^43,44^ Other measures of variability include the coefficient of variation (CoV), average real variability (ARV), and the variability independent of the mean (VIM).^15^ CoV, the ratio of SD to the mean, is more sensitive to lower BP values and is used as a method to mitigate the influence of mean arterial BP on mortality.^45^ ARV, derived from the differences between consecutive BP readings and averaged by dividing the sum by the number of measurements minus one, has shown promise as a superior measure of BPV.^46,47^ While shorter durations of ARV provide valuable insights with continuous beat-by-beat monitoring,^48^ full 24-hour monitoring provides a more comprehensive assessment of daily BPV outside of the laboratory. VIM measures variability uncorrelated with mean BP by adjusting the SD of BP with the power of the mean.^49^ We calculated these four BPV indices to provide a comprehensive profile of variability, as each measure captures different statistical and physiological aspects of BP fluctuations. While ARV is widely used in ABPM research for its ability to quantify changes between consecutive readings,^50^ it does not fully capture all variability patterns relevant to SCI. Therefore, we did not designate a single “primary” BPV outcome, but for the purposes of this manuscript, considered these four indices equally important and complementary. This multi-index approach provides a more comprehensive clinical interpretation and cross-study comparability.

### Nocturnal Dip

As individuals with chronic SCI often experience sleep-onset BP perturbations due to transfers, care routines, and dysreflexic episodes,^51^ we evaluated nocturnal dip for SBP and DBP by first averaging the lowest nighttime SBP or DBP values with the two nearest respective nighttime readings. This average was then subtracted from the average median SBP and DBP values obtained during the day. The percentage was determined by dividing the difference between these average nighttime readings and the average median daytime BP by the average median daytime BP, then multiplying by 100. Participants were sub-classified according to the percentage of nighttime BP decline as follows: extreme dippers if nighttime BP reduction ≥20%; dippers if the decline was between 10-20%; non-dippers if the decline was between 0-10%; and the reverse dippers if there was an increase in nighttime BP.^52^

### Blood Pressure Fluctuations

Individuals with SCI at or above T6 frequently experience AD and hypotensive episodes,^8,10,53,54^ leading to pronounced BP fluctuations that contribute to increased BPV. We classified hypotensive events as SBP <100 mmHg and DBP <70 mmHg,^55^ excluding nighttime BP (to avoid misclassifying nocturnal dip as hypotensive events). Because individuals with chronic SCI commonly have resting BP values below traditional clinical cut-offs, these absolute thresholds were applied to quantify hypotensive events in a manner appropriate for the SCI population, rather than to diagnose hypotension based on general-population criteria. To differentiate persistent hypotension from discrete hypotensive episodes, we calculated for each participant the percentage of total 24-hour readings within the hypotensive range and their median 24-hour SBP and DBP. Participants with more than 50% of readings below threshold were classified as persistently hypotensive, whereas those with fewer were considered to exhibit episodic events. This distinction allowed us to separate individuals with chronically low BP profiles from those experiencing transient drops related to autonomic instability. Hypertensive events were defined as SBP >150 mmHg, reflecting a substantial deviation from typical resting SBP values in individuals with C-SCI, which often range from 90-110 mmHg.^56^ This threshold aligns with the value recommended by the Consortium of Spinal Cord Medicine for pharmacological management of a refractory episode of AD.^57^ Although no universally accepted BP threshold defines immediate danger in SCI, the use of absolute cutoffs is consistent with existing clinical guidelines and commonly used criteria. This allows for consistent classification of BP events across participants and captures the extreme fluctuations that contribute to increased BPV.^58^

### Statistical Analysis

Data are expressed as mean ± standard deviation. Demographics, mean of the median SBP, mean of the median DBP, along with BPV metrics for SBP and DBP (i.e., SD, CoV, ARV, and VIM) were summarized and statistically compared between groups using analysis of variance (ANOVA) with Bonferroni post hoc (to account for different types of variables) or Kruskal Wallis with Mann-Whitney U post hoc, if non-parametric. Although ANOVA is conventionally used to compare group means, in this study it was applied to BPV indices (e.g., ARV, SD, CoV, VIM), comparing the average BPV index values between groups, rather than the raw variance of BP readings, allowing group-level differences in BPV indices to be tested statistically. In addition to Bonferroni-adjusted post hoc tests, false discovery rate (FDR) control using the Benjamini–Hochberg procedure (α = 0.05) was applied separately across the four systolic and four diastolic BPV indices to account for multiple testing across indices. Eta squared (η^2^) was calculated to determine effect size and was defined as small (η^2^ ≥ 0.01), medium (η^2^ ≥ 0.06), and large (η^2^ ≥ 0.14) effects.^59^ Tests were two-sided with a significance level of 0.05, unless otherwise specified. Statistical analyses were performed using standard statistical packages in R (Version 4.4.1, R Foundation for Statistical Computing, Vienna, Austria). Graphical representations were created in R Studio and Adobe Illustrator (Version 28.1, Adobe Inc., 2023, San Jose, CA, USA).

BPV was assessed using SD (mean difference from mean BP), CoV (SD divided by mean), ARV (mean of absolute successive differences), and VIM (adjustment of SD using a power transformation of the mean BP). The prevalence of SBP and DBP dipping patterns (i.e., extreme dippers, dippers, non-dippers, and reverse dippers)^28,29^ were compared across the C-SCI, UT-SCI, and NI-C groups using chi-square (□^2^) tests.

The relationship between BP lability and NLI was examined using a negative binomial regression analysis. The prevalence of hypotensive and hypertensive events was assessed by using chi-square tests on the participants in each group. We further examined the distributional characteristics of the ABPM data by calculating the skewness and kurtosis of each participant’s ABPM recordings to further describe BP differences between groups. Skewness indicates the symmetry of a distribution, with zero showing perfect symmetry, while kurtosis measures the relative heaviness of the tails of a distribution, with higher values indicating heavier tails and greater departure from normality, rather than necessarily more outliers. Negatively skewed distributions have tails toward lower values, and leptokurtic distributions have thicker tails and increased BP lability.^55^ The z-scores for skewness and kurtosis were also calculated to characterize the magnitude of the deviations from normality for each participant by group.^60^

An unbiased recursive partitioning conditioning inference tree (URP-CTREE), a regression model that conducts sequential tests of independence among predictors (biological sex, age, Time Since Injury (TSI), weight, NLI, AIS, and each research study), was utilized to explore the link between these predictors and 24-hour ABPM indices. A more detailed description of URP-CTREE can be found here.^61^

To assess the robustness of our pooled analysis, we performed sensitivity analyses by examining the SCI cohorts between the four studies. Specifically, we conducted one-way ANOVA tests to evaluate the impact of the sub-studies on SBP of SD, CoV, ARV, and VIM. Post hoc pairwise comparisons were conducted using Tukey’s HSD test (for Type II error control and more power) to explore significant differences between subgroups.

As BP was recorded every 15 minutes during the daytime and at least hourly overnight, we evaluated whether unequal sampling density biased BPV estimates. To test this, we performed a sensitivity analysis in which daytime BP data were “hourly-thinned” by aggregating all readings within each hour to their median SBP value. Participants were included if their number of retained daytime hourly bins was at least equal to their number of valid nighttime hourly bins. BPV indices (SD, CoV, ARV, and VIM for SBP) were recalculated using this reduced dataset. This conservative approach further ensured that variability estimates were not inflated by clusters irregularly spaced variability measurements (e.g., from symptom-triggered readings). We note that previous work has discussed the importance of consistent measurement intervals and sampling frequency in BPV research,^62,63^ however we attempted to account for the known acute, episodic BP surges and drops common in SCI, which can produce clusters of readings and artificially elevate time-domain BPV metrics. To our knowledge no standard protocol describes “hourly-thinning” as we have done to account for this unequal sampling. Therefore, this procedure is presented as a supplemental sensitivity analysis, and report the results alongside the primary dataset.

## Results

Data were collected from 80 participants (STARD Flow Diagram; Supplementary Figure S1), including 44 with C-SCI, 19 with UT-SCI, and 17 NI-C. The cohort was predominantly male (80%). The measurement ratio, defined as the number of successful brachial BP recordings out of the pre-programmed attempts, showed no significant differences among the groups. Likewise, the total number of measurements taken over 24 hours did not differ significantly. The groups were also comparable in terms of age, weight, and time since injury (TSI; for participants with SCI). All data are summarized in Table 1.

**Table 1.** Participant Characteristics.

| Outcome | C-SCI (n=44) | UT-SCI (n=19) | NI-C (n=17) | P value |
| --- | --- | --- | --- | --- |
| Age [years], mean ± SD | 43 ± 10 | 43 ± 13 | 41 ± 15 | 0.77 |
| Sex (M), n (%) | 34 (77%) | 16 (84%) | 14 (82%) | 0.79 |
| Body mass [Kg] | 75 ± 20 | 77 ± 14 | 82 ± 13 | 0.42 |
| NLI | C1 = 1 | T2 = 3 |  |  |
|  | C4 = 8 | T3 = 5 |  |  |
|  | C5 = 19 | T4 = 6 | - | - |
|  | C6 = 11 | T5 = 4 |  |  |
|  | C7 = 4 | T6 = 1 |  |  |
|  | C8 = 1 |  |  |  |
| AIS Grade | A = 20 |  |  |  |
|  | B = 18 | A = 18 |  |  |
|  | C = 4 | B = 1 | - | - |
|  | D = 2 |  |  |  |
| TSI [years] | 15 ± 9 | 17 ± 14 | - | 0.51 |
| Measurement Ratio | 87 ± 12 | 86 ± 18 | 86 ± 14 | 0.95 |
| Number of Successful Measurements | 67 ± 17 | 66 ± 23 | 65 ± 15 | 0.93 |

Abbreviations: A, complete injury (no sensory/motor function preserved); AIS, American Spinal Injury Association (ASIA) Impairment Scale; B, sensory incomplete; C, motor incomplete with >50% key muscles below neurological level of injury having no active movement against gravity or resistance; C-SCI, cervical spinal cord injury; C1-C8, cervical spinal cord segments 1-8; D, motor incomplete with ≥ 50% key muscles below neurological level of injury having at least active movement against gravity but not necessarily resistance; F, female; M, male; NI-C, non-injured control; NLI, neurological level of injury; T2-T6, thoracic spinal cord segments 2-6; TSI, time since injury; UT-SCI, upper-thoracic spinal cord injury

### Blood Pressure Variability Indices

Standard deviation for SBP was significantly (*P* = 0.004, η^2^ = 0.130) different across groups, being significantly higher in the C-SCI group (18 ± 5 mmHg) compared to the UT-SCI (14 ± 4 mmHg) and NI-C (14 ± 3 mmHg) groups. CoV for SBP was significantly (*P* < 0.001, η^2^ = 0.227) different across groups, being significantly higher in the C-SCI group (16 ± 5 %) compared to the UT-SCI (12 ± 3 %) and NI-C (11 ± 2 %) groups. ARV for SBP was significantly (*P* < 0.001, η^2^ = 0.209) different across groups, being significantly higher in the C-SCI group (13 ± 4 mmHg) compared to the UT-SCI (11 ± 2 mmHg) and NI-C (10 ± 2 mmHg) group. VIM for SBP was significantly (*P* = 0.002, η^2^ = 0.154) different across groups, being significantly higher in the C-SCI group (17 ± 5 mmHg) compared to the UT-SCI (14 ± 4 mmHg) and NI-C (13 ± 2 mmHg) group (Figure 1).

**Figure 1.**
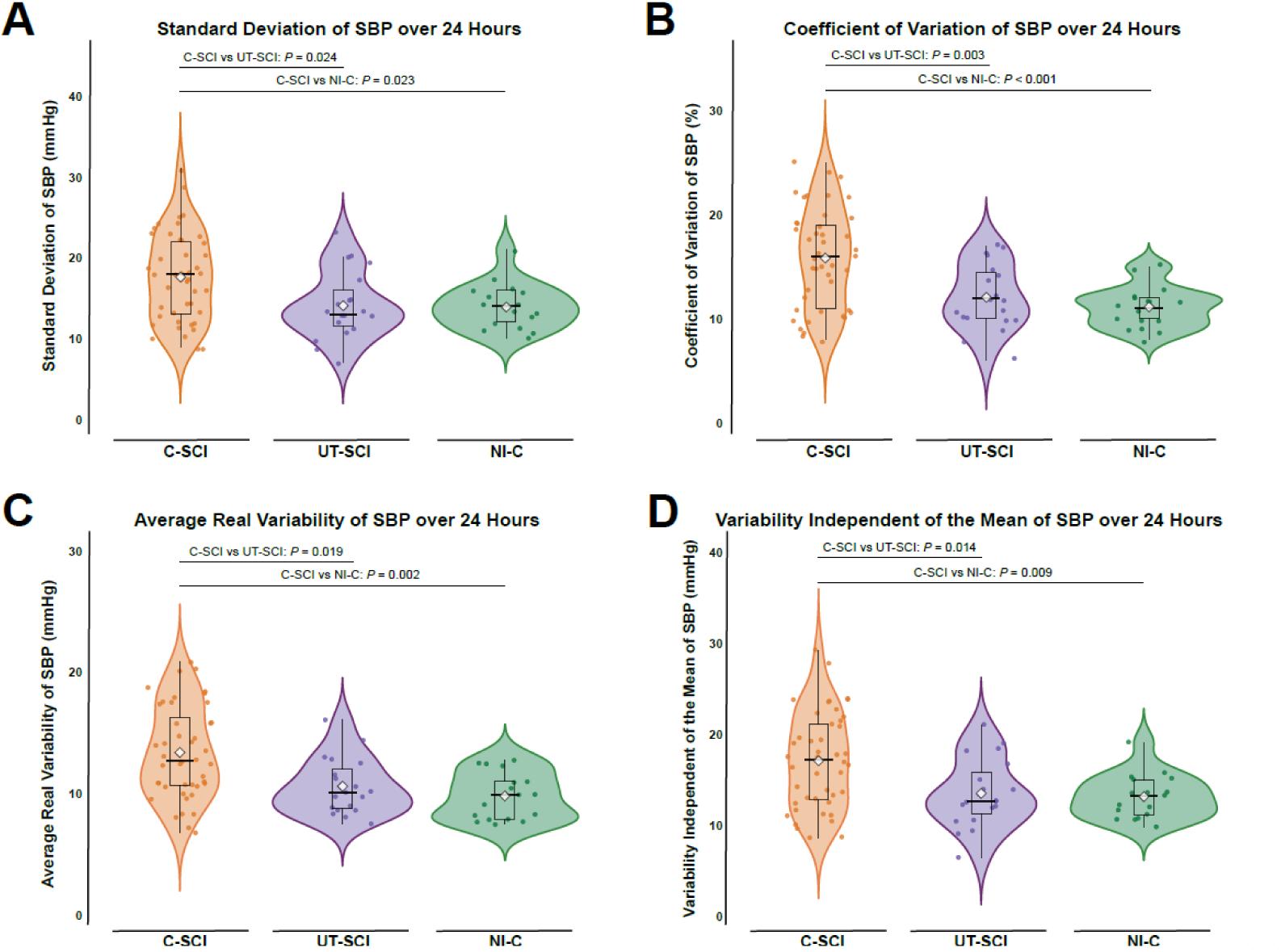
Comparison of systolic blood pressure variability indices over a 24-hour period. **(A)** Standard deviation (SD) for systolic blood pressure (SBP) over 24-hours, **(B)** Coefficient of Variation (CoV) for SBP over 24-hours, **(C)** Average Real Variability (ARV) for SBP over 24-hours, **(D)** Variability Independent of the Mean (VIM) for SBP over 24-hours. Violin plots are presented with individual participant data points with box-and-whisker plots overlayed; the thick horizonal black line represents the median value of the respective variable and the white diamond represents the mean. Orange represents the cervical spinal cord injury (C-SCI) group, purple represents the upper-thoracic spinal cord injury (UT-SCI) group, and green represents the non-injured control group (NI-C). Statistical significance using a Bonferroni-adjusted post hoc test is shown if the one-way ANOVA (normal distribution) or Kruskal-Wallis (not normal distribution) for comparison across all groups was significant. See Supplementary Data for detailed statistics.

Effect sizes indicated large effects of neurological level of injury on systolic BPV indices (η² = 0.27-0.34). After FDR correction across indices, all four systolic indices (SD, CoV, ARV, and VIM) remained significant (q ≤ 0.002). For diastolic BPV, only CoV remained significant after FDR correction (q = 0.0496; η² = 0.12).

### Median SBP and DBP and Nocturnal Dip

The mean of the median SBP over 24 hours was significantly (*P* < 0.001, η^2^ = 0.268) different across groups, being significantly higher in the NI-C group (127 ± 10 mmHg) compared to the C-SCI (109 ± 13 mmHg) and UT-SCI (115 ± 12 mmHg) group. The mean of the median DBP over 24 hours was significantly (*P* < 0.001, η^2^ = 0.259) different across groups, being significantly higher in the NI-C group (78 ± 10 mmHg) compared to the C-SCI (65 ± 9 mmHg) and UT-SCI (67 ± 7 mmHg) group. Nocturnal SBP dip was significantly (*P* < 0.001, η^2^ = 0.221) different across groups, being significantly lower in the C-SCI group (-6 ± 12%) compared to the UT-SCI (-17 ± 8%) and NI-C (-18 ± 7%) group. Nocturnal DBP dip was significantly (*P* < 0.001, η^2^ = 0.262) different across groups, being significantly lower in the C-SCI group (-10 ± 14%) compared to the UT-SCI (-23 ± 11%) and NI-C (-26 ± 6%) group (Figure 2). Moreover, a greater proportion of reverse dippers were found in the C-SCI group compared to the UT-SCI or NI-C groups for both SBP and DBP (Supplementary Figure S2).

**Figure 2.**
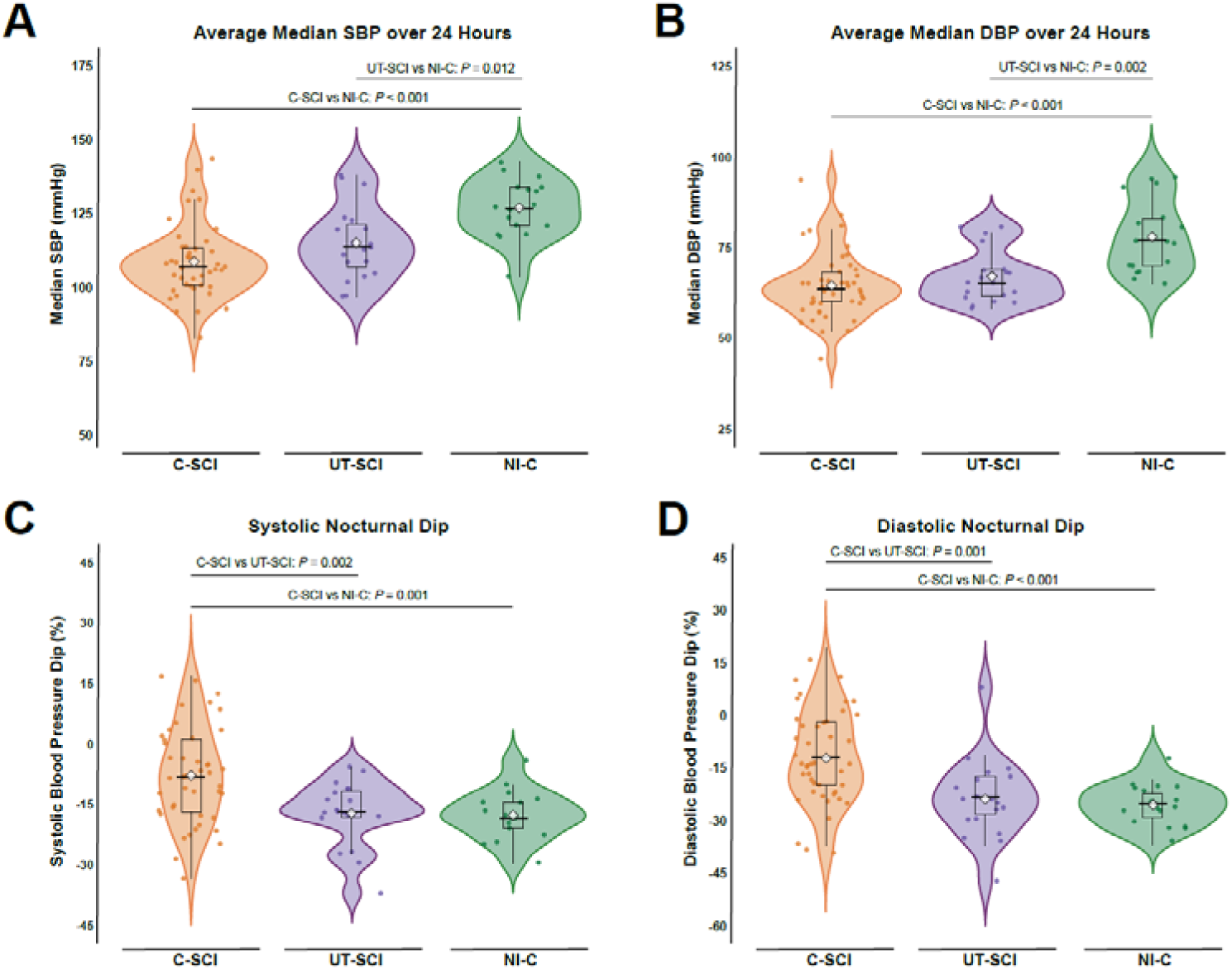
Comparison of median blood pressure readings over a 24-hour period with assessment of nocturnal dip. **(A)** Mean of the median systolic blood pressure (SBP) over 24-hours, **(B)** Median diastolic blood pressure (DBP) over 24-hours, **(C)** Systolic blood pressure nocturnal dip over 24-hours, **(D)** Diastolic blood pressure nocturnal dip over 24-hours. Violin plots are presented with individual participant data points with box-and-whisker plots overlayed; the thick horizonal black line represents the median value of the respective variable and the white diamond represents the mean. Orange represents the cervical spinal cord injury (C-SCI) group, purple represents the upper-thoracic spinal cord injury (UT-SCI) group, and green represents the non-injured control group (NI-C). Statistical significance using a Bonferroni-adjusted post hoc test is shown if the one-way ANOVA (normal distribution) or Kruskal-Wallis (not normal distribution) for comparison across all groups was significant. See Supplementary Data for detailed statistics.

### Blood Pressure Lability

The percentage of brachial BP recordings that met the criteria for hypotensive or hypertensive events are shown in Figure 3A and 3B, respectively, to account for the different number of measurements across participants. A negative binomial regression analyses in a hierarchical structure was performed to identify significant contributors to BP lability with NI-C serving as the reference category. The primary dependent variable was BP lability, the total number of recordings for each participant over the 24-hour period was a covariate, and NLI was the categorical independent variable. For hypotensive events, both C-SCI (^β = 3.072, *P* < 0.001) and UT-SCI (^β = 2.319, *P* < 0.001) were strongly associated with a higher count of hypotensive events compared to NI-C. When exponentiating the extracted coefficient, C-SCI had a multiplicative effect of 21.6 and UT-SCI had a multiplicative effect of 10.2, corresponding to a substantial increase in the number of hypotensive events, especially in those with C-SCI. For hypertensive events, NLI showed a significant decrease for UT-SCI (^β = -0.304, *P* < 0.001), while the association with C-SCI was not statistically significant (^β = -0.097, *P* = 0.12). The number of hypotensive and hypertensive events can be found in Supplementary Table S2.

**Figure 3.**
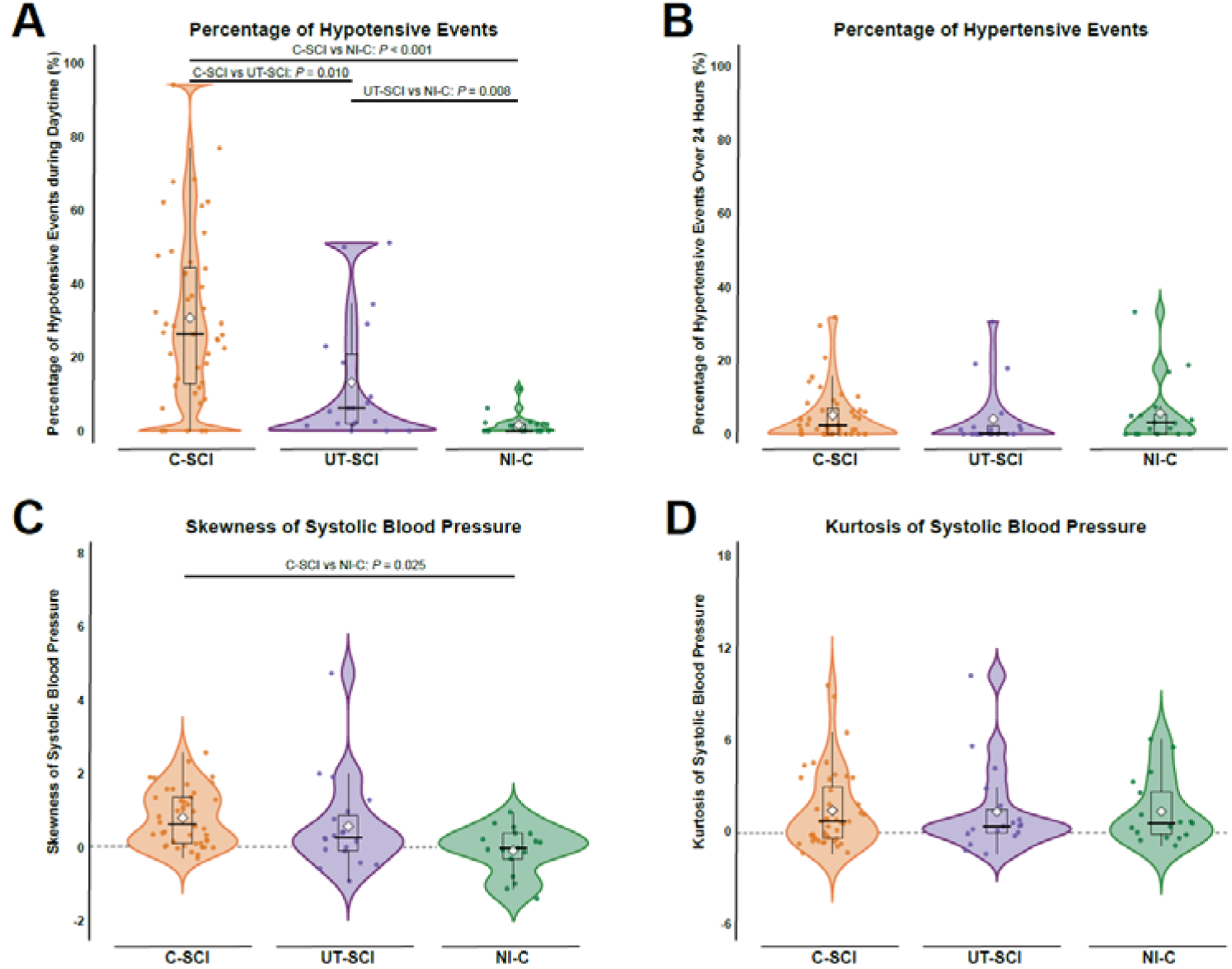
Comparing hypotensive and hypertensive events across different injury characteristics. **(A)** The percentage of hypotensive events for each participant over the 24-hour ABPM recording period categorized by neurological level of injury (NLI), **(B)** The percentage of hypertensive events for each participant over the 24-hour ABPM recording period categorized by NLI, **(C)** The skewness of the SBP values were calculated for each participant independently, **(D)** The kurtosis of the SBP values were calculated for each participant independently. Violin plots are presented with individual participant data points with box-and-whisker plots overlayed; the thick horizonal black line represents the median value of the respective variable and the white diamond represents the mean. Orange represents the cervical spinal cord injury (C-SCI) group, purple represents the upper-thoracic spinal cord injury (UT-SCI) group, and green represents the non-injured control group (NI-C). Statistical significance using a Bonferroni-adjusted post hoc test is shown if the one-way ANOVA (normal distribution) or Kruskal-Wallis (not normal distribution) for comparison across all groups was significant. See Supplementary Data for detailed statistics.

For hypotension, the effect of NLI was significant (χ² = 32.514, P < 0.001), with the highest prevalence observed in the C-SCI group. The mean percentage of hypotensive readings was highest in the C-SCI group (31.0%), followed by UT-SCI (17.8%), and NI-C (4.7%). Several individuals with C-SCI and UT-SCI had >40% of readings within this range, consistent with persistent hypotension, while others exhibited only occasional low-BP episodes (< 20%). Therefore, individuals with higher-level SCI exhibit a substantial burden of low BP across the 24-hour period.

The SBP skewness distribution (Figure 3C) differed significantly across groups (P = 0.004, η² = 0.136), being markedly right-skewed in C-SCI (0.80 ± 0.76) and moderately skewed in UT-SCI (0.58 ± 1.27) compared with a near-symmetrical NI-C distribution (-0.08 ± 0.65). SBP kurtosis (Figure 3D) did not differ significantly between groups (P = 0.10, η² = 0.023). Z-score analysis further revealed that 50% of the C-SCI group and 42% of the UT-SCI group exhibited significantly non-Gaussian SBP distributions, compared with 35% of NI-C. These deviations from normality likely reflect the coexistence of persistent hypotension and episodic BP surges characteristic of autonomic dysregulation after high-level SCI.

The URP-CTREE analysis was used to identify subgroups by NLI (C-SCI, UT-SCI, NI-C), TSI, age, sex, and study identifier to assess systolic BPV indices (SD, CoV, ARV, VIM) over 24 hours. NLI emerged as the strongest predictor, accounting for a moderate proportion of the variance in BPV (R^2^: 0.26-0.32). Conditional inference trees illustrating the BPV trends are provided in Supplementary Figure S3.

### Sensitivity Analysis

The one-way ANOVA for SBP did not reveal significant differences between the SCI trials for SD (*P* = 0.49), CoV (*P* = 0.57), ARV (*P* = 0.19), and VIM (*P* = 0.17). No significant differences were found for SD, CoV, ARV, or VIM, supports the validity of pooling these datasets despite minor differences in the eligibility criteria of the original study protocols. Sensitivity analyses using the hourly-thinned daytime dataset (median within hour) produced qualitatively similar results to the full 24-hour dataset (Supplementary Table S3). This indicates that the elevated BPV observed in C-SCI is not an artifact of unequal daytime/nighttime sampling frequency.

## Discussion

Our results demonstrate significant alterations in ambulatory BP components among individuals with SCI, particularly those with C-SCI, highlighting notable cardiovascular dysregulation in this population (*the central finding of this study*). Elevated BPV, a known predictor of cardiovascular morbidity and mortality in non-SCI populations,^64^ warrants investigation as a potential prognostic marker in individuals with SCI. Although our findings reveal clear alterations in BP components, longitudinal studies are required to determine whether these BPV abnormalities translate into increased cardiovascular risk in this population.

### Blood Pressure Variability

Time-domain BPV indices, including SD, CoV, ARV, and VIM, were significantly elevated in the C-SCI group compared to UT-SCI and NI-C. Complementary analyses, such as unbiased recursive conditional inference trees (Supplementary Figure S3), identified NLI as the primary determinant of BPV, independent of other factors such as TSI or AIS severity. Elevated BPV in individuals with C-SCI likely reflects impaired autonomic cardiovascular regulation,^65^ emphasizing the need for targeted BP management strategies in this specific group. Unlike the current focus of achieving normotensive BP targets,^66,67^ our findings suggest that reducing BPV, particularly in those with high-level chronic SCI, may also be important as previous work has linked higher BPV to increased risk of cardiovascular events.^68^

A study of over 42,000 non-SCI adults reported that BPV reliably predicts cardiovascular outcomes and is associated with increased risk for new onset of CVD, such as myocardial infarction, stroke, and heart failure.^69^ This association persisted across various age groups, sexes, and clinical comorbidities. Our data further indicates that individuals with C-SCI exhibit a much wider range of BPV compared to NI-C, characterized by more pronounced BP fluctuations and the absence of a normal nocturnal dip in BP.

Despite the increasing interest in BPV metrics, few studies have explored these measures in the SCI population. Consistent with our findings, another study found that individuals with C-SCI (n = 33) and UT-SCI (n = 3) experience significant BP instability, showing frequent asymptomatic fluctuations between hypertension and hypotension episodes compared to ambulatory NI-C (n = 13).^14^ In terms of CoV for SBP during the day (awake), our data (Supplementary Table S2) reported 16 ± 5% for C-SCI, 11 ± 3% for UT-SCI, and 10 ± 2% in NI-C (*P* < 0.001), similar to the values of 15 ± 1% for SCI versus 10 ± 1% for NI-C (*P* = 0.0013) reported by Wang and colleagues.^14^ ARV for SBP during the day (awake) also mirrored these findings, with our study showing 14 ± 4mmHg for C-SCI, 11 ± 3mmHg for UT-SCI, and 10 ± 2mmHg for NI-C (*P* < 0.001), compared to 13 ± 1mmHg for SCI and 10 ± 1mmHg for NI-C. Our study’s use of data from multiple studies, resulting in a larger number of UT-SCI participants, allowed for a more thorough stratification by NLI, providing more insight into its role in cardiovascular dysregulation, which was not addressed in the previous study.^14^ Additionally, while the other study focused on fewer BPV indices, our study examined both SD and VIM, offering for the first time a more comprehensive evaluation of individuals with SCI.

The extent to which mechanisms underlying elevated BPV in individuals with SCI mirror those observed in non-injured individuals with cardiovascular disease remains poorly understood. In non-SCI populations, elevated BPV has been associated with arterial stiffness, impaired baroreflex sensitivity, endothelial dysfunction, and systemic inflammation, each of which has been implicated in long-term cardiovascular risk. Importantly, many of these abnormalities have also been independently documented in individuals with SCI.^70–73^ However, in contrast to non-SCI populations, elevated BPV in cervical SCI (C-SCI) is likely dominated by injury-related autonomic dysregulation, rather than vascular or endothelial factors, with episodic sympathetic surges such as AD playing a key role.

Systolic BPV, which is linked to arterial stiffness and aging in non-SCI populations,^74^ may be particularly relevant for individuals with SCI, given that this condition is often described as a model of premature aging.^70,72,75^ Evidence indicates that individuals with SCI, especially those with paraplegia, exhibit higher aortic pulse wave velocity (aPWV; a marker of arterial stiffness), compared to individuals with tetraplegia,^76^ although lower resting BP in tetraplegia may partially influence these findings.^74^ Diastolic BPV (see Supplementary Table S2), which has been associated with autonomic and endothelial dysfunction,^74^ is also exaggerated in the SCI population.^77^ Our prior work further supports the presence of persistent central arterial stiffness in SCI, even when individuals are well matched with NI-C.^72^ Together, these observations suggest that BPV in SCI arises from the interaction between autonomic impairment and vascular remodelling. Longitudinal studies are required to clarify the role of BPV in risk stratification, whether therapeutic interventions (such as exercise ^78^) can modulate BPV and to inform targeted interventions aimed at reducing cardiovascular risk in SCI more generally.

### Blunted Nocturnal Dipping

Individuals with C-SCI demonstrated a markedly attenuated nocturnal dip compared to UT-SCI and NI-C. Consistent with prior observations in SCI,^79,80^ many participants with C-SCI exhibited non-dipping or reverse-dipping patterns, reflecting substantial disruption of autonomic cardiovascular regulation in this group. In contrast, UT-SCI participants showed more preserved, though still variable, nocturnal BP declines.

Although abnormal nocturnal BP patterns (for non-dipping and reverse dipping) have been associated in the general population with adverse cardiovascular markers such as stroke risk, subclinical organ damage, and mortality,^81–86^ our study did not assess cardiovascular outcomes or intermediate CVD biomarkers. Furthermore, nocturnal dipping patterns should be reported in the context of overall BP level. In individuals with chronic SCI who are persistently hypotensive, nocturnal dip does not necessarily reflect physiological cardiovascular recovery during sleep, as it might in normotensive or hypertensive populations. For example, a 20% nighttime BP reduction in a participant with a mean SBP of 90 mmHg would represent profound hypotension rather than a healthy circadian rhythm. Thus, conventional dipper/non-dipper classifications may not accurately capture cardiovascular health in individuals with chronically low baseline BP, and alternative or adjusted interpretative frameworks may be warranted in this population.

Nevertheless, given the established associations between circadian BP disruption and cardiovascular vulnerability in other populations,^82,85–88^ the high prevalence of blunted or reversed nocturnal dipping in C-SCI emphasizes the degree of autonomic impairment in this group. Whether restoring more physiological nocturnal BP patterns could improve cardiovascular health in SCI remains unknown and warrants further investigation. These findings should be viewed as descriptive evidence of altered BP regulation rather than as clinical prognostic indicators.

### Altered Blood Pressure Dynamics in SCI

Participants with C-SCI and UT-SCI exhibited profound BP regulation impairments, characterized by a higher frequency of hypotensive events compared to NI-C. The mean proportion of hypotensive readings was over six-fold higher in C-SCI than in controls (31% vs 5%). Several individuals with C-SCI and UT-SCI had more than 40% of readings in this range, indicative of persistent hypotension rather than isolated events. This pattern likely reflects impaired sympathetic vasoconstrictor control and blunted baroreflex buffering following disruption of descending sympathetic pathways.^89^ Persistent hypotension may exacerbate the degree of orthostatic, postprandial, and post-exercise hypotension in individuals with SCI, thereby resulting in chronic end-organ hypoperfusion.^90^ Given the lower resting BP commonly observed in SCI, even modest additional BP reductions may be sufficient to cross cerebral perfusion thresholds and provoke symptoms,^91^ whereas comparable relative BP reductions in individuals with higher baseline BP may remain asymptomatic.

Although hypertensive events were not directly associated with NLI in our study, they remain important to monitor, particularly to capture transient spikes such as those caused by AD.^16^ Elevated SBP increases cardiovascular workload and accelerates arterial damage, leading to an increased myocardial infarction and stroke risk.^92^ Since SBP is a more reliable predictor of future cardiovascular risk than DBP,^93,94^ targeting systolic BPV indices may be more important in mitigating cardiovascular risks in the SCI population.

In addition to event frequency, our analysis of distributional characteristics revealed that BP patterns in SCI are often non-Gaussian (not normal) in shape, particularly in the C-SCI group. Using a z-score-based detection of skewness and kurtosis, we found 50% of C-SCI and 42% of UT-SCI participants in our cohort demonstrated significantly skewed or heavy-tailed SBP distributions, indicating a higher prevalence of extreme SBP values and BP lability. Such distributional abnormalities may represent greater end-organ stress, as repeated transient spikes or drops in BP can damage the vascular endothelium, even when mean BP appears within normal limits.^95^ These findings align with previous work showing increased skewed SBP distributions in individuals with tetraplegia and non-Gaussian profiles in upper-thoracic injuries.^55^ Therefore, z-score-based detection may also improve individualized BP monitoring as it is sensitive to identifying BP instability that may not be captured by daily mean or median BP values alone.

### Clinical Considerations and Future Directions

From a practical standpoint, these findings support the routine use of 24-hour ABPM in individuals with high-level SCI to capture BP fluctuations that are frequently not captured during a typical clinic visit with the current standard of averaging multiple blood pressures. Clinicians should interpret BP patterns beyond mean values, paying attention to variability indices and nocturnal dipping profiles. Identifying individuals with exaggerated BPV or reverse dipping may help guide individualized management, though standardized thresholds for BPV in SCI are not established. BPV impacts vascular endothelial cells via altered wall shear stress, exacerbating atherosclerotic plaque formations.^96^ This accelerated plaque formation occurs though molecular pathways, including ion channel activation and DNA changes. Proteins like Piezo1, integral to cardiovascular regulation, are also affected, promoting plaque development via the NF-кB pathway.^97^ These insights emphasize the importance of stabilizing BPV as a therapeutic target to reduce atherosclerosis progression.

While our study highlights the role of BPV in cardiovascular risk, the inclusion of concurrent arterial stiffness assessments or inflammatory markers would have strengthened our findings, allowing for a more direct link between BPV and established prognostic risk factors, facilitating a more comprehensive CVD risk stratification. Addressing BPV through comprehensive assessments, including 24-hour ABPM, may improve cardiovascular risk stratification and management for these individuals.

Although unnoticed by many individuals with SCI,^14^ SBP variability and arterial stiffness is more significantly correlated to the risk of stroke^98^ and cognitive decline.^99,100^ Exaggerated cardiovascular responses to stimuli in SCI, such as urinary bladder distension or postural changes, reflect impaired baroreceptor reflexes and increased sensitivity to vasopressin and noradrenaline.^101^ Our findings emphasize the need for BPV and altered BP regulation monitoring in individuals living with SCI, especially for high-level injuries.

Consistent with prior research, lesion-dependent cardiovascular impairments necessitate a comprehensive assessment beyond mean 24-hour BP values, incorporating BPV and nocturnal dipping patterns.^76^ While 24-hour ABPM is ideal, other BPV measurement methods, such as home monitoring (e.g., self-measured BP readings) and clinic measurements (e.g., standardized in-office readings), also predict cardiovascular outcomes, though these indices are not interchangeable.^34^ Increased BPV independently predicts cardiovascular morbidity and mortality,^102,103^ with all BPV indices showing prognostic value, despite moderate inter-index correlations.^104–107^ Addressing autonomic dysfunction and targeting BPV stability via tailored interventions may reduce cardiovascular risks in the SCI population, warranting further studies investigating the effect of therapeutic strategies.^105,107^

### Limitations

Despite its strengths, our study has a few considerations. While data was mainly collected from a sample representative of the SCI population (∼80% male) with the most severe cardiovascular consequences, these characteristics may limit the generalizability to females with SCI. Additionally, the data were merged from different studies, which may introduce some variability; however, a sensitivity analysis was performed to demonstrate the consistency of the findings. Moreover, our study focused on a 24-hour ABPM period, which provides valuable information on diurnal BP patterns, but does not account for the longer-term variations and seasonal influences that could affect BP and autonomic function.^108^ Unmeasured factors such as lifestyle (e.g., physical activity levels), medication use, and psychological stressors were not measured and should be considered in future studies. Finally, two ABPM devices (Mobil-O-Graph and Meditech card(X)plore) were used across sub-studies. Although both devices have been validated for brachial BP measurement according to international protocols,^40^ device-level assignment per participant was not retained, precluding device-stratified analyses. While time-domain BPV indices (SD, CoV, ARV, VIM) are unaffected by constant calibration differences and our standardized sampling/processing reduces device-related variation, differences in intrinsic measurement properties could still influence BPV values. Additionally, our between-study sensitivity analyses and conditional inference trees pointed to NLI as the primary determinant of BPV; if device type had been a dominant driver, we would expect inconsistent effects by study (which were not observed).

### Conclusion

Our findings emphasize the significant impact of SCI, particularly C-SCI, on BPV, cardiovascular regulation, and nocturnal dipping. Individuals with C-SCI exhibited greater BP instability and a higher burden of persistent hypotension, reflecting impaired sympathetic control. These alterations likely contribute to the elevated cardiovascular morbidity observed in this population. Collectively, the results highlight the necessity for targeted cardiovascular monitoring and intervention strategies in this vulnerable population. Further research is needed to explore the underlying mechanisms, clinical implications, and therapeutic approaches for managing BP dysregulation in individuals living with SCI.

## Data Availability

Data available upon reasonable request.

## Acknowledgements

We thank the participants for taking part in the study.

---

**Supplementary Table 1.**
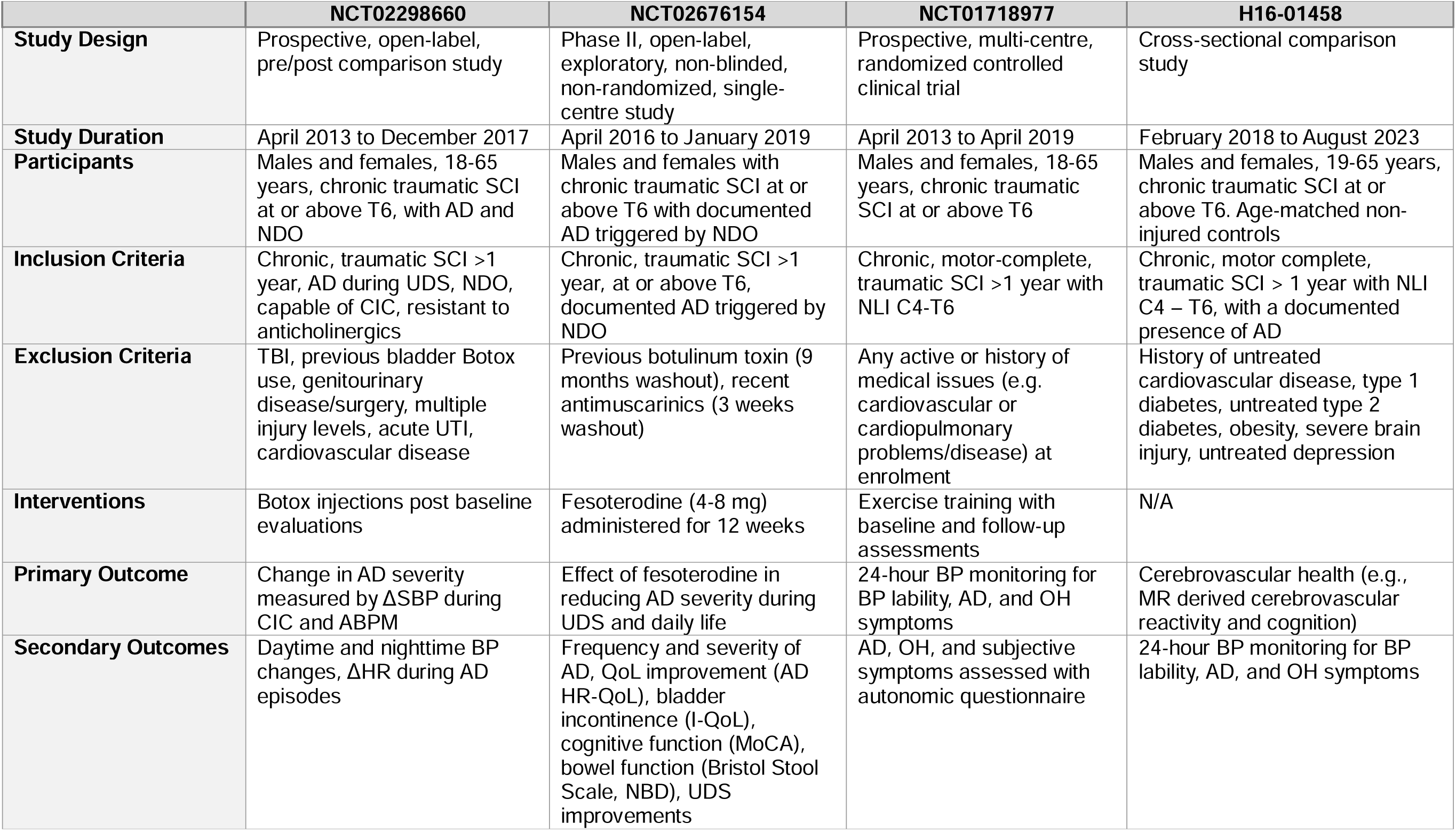

|  | **NCT02298660** | **NCT02676154** | **NCT01718977** | **H16-01458** |
| --- | --- | --- | --- | --- |
| **Study Design** | Prospective, open-label, pre/post comparison study | Phase II, open-label, exploratory, non-blinded, non-randomized, single-centre study | Prospective, multi-centre, randomized controlled clinical trial | Cross-sectional comparison study |
| **Study Duration** | April 2013 to December 2017 | April 2016 to January 2019 | April 2013 to April 2019 | February 2018 to August 2023 |
| **Participants** | Males and females, 18-65 years, chronic traumatic SCI at or above T6, with AD and NDO | Males and females with chronic traumatic SCI at or above T6 with documented AD triggered by NDO | Males and females, 18-65 years, chronic traumatic SCI at or above T6 | Males and females, 19-65 years, chronic traumatic SCI at or above T6. Age-matched non-injured controls |
| **Inclusion Criteria** | Chronic, traumatic SCI >1 year, AD during UDS, NDO, capable of CIC, resistant to anticholinergics | Chronic, traumatic SCI >1 year, at or above T6, documented AD triggered by NDO | Chronic, motor-complete, traumatic SCI >1 year with NLI C4-T6 | Chronic, motor complete, traumatic SCI > 1 year with NLI C4 – T6, with a documented presence of AD |
| **Exclusion Criteria** | TBI, previous bladder Botox use, genitourinary disease/surgery, multiple injury levels, acute UTI, cardiovascular disease | Previous botulinum toxin (9 months washout), recent antimuscarinics (3 weeks washout) | Any active or history of medical issues (e.g. cardiovascular or cardiopulmonary problems/disease) at enrolment | History of untreated cardiovascular disease, type 1 diabetes, untreated type 2 diabetes, obesity, severe brain injury, untreated depression |
| **Interventions** | Botox injections post baseline evaluations | Fesoterodine (4-8 mg) administered for 12 weeks | Exercise training with baseline and follow-up assessments | N/A |
| **Primary Outcome** | Change in AD severity measured by ΔSBP during CIC and ABPM | Effect of fesoterodine in reducing AD severity during UDS and daily life | 24-hour BP monitoring for BP lability, AD, and OH symptoms | Cerebrovascular health (e.g., MR derived cerebrovascular reactivity and cognition) |
| **Secondary Outcomes** | Daytime and nighttime BP changes, ΔHR during AD episodes | Frequency and severity of AD, QoL improvement (AD HR-QoL), bladder incontinence (I-QoL), cognitive function (MoCA), bowel function (Bristol Stool Scale, NBD), UDS improvements | AD, OH, and subjective symptoms assessed with autonomic questionnaire | 24-hour BP monitoring for BP lability, AD, and OH symptoms |

**Supplementary Table 2.**
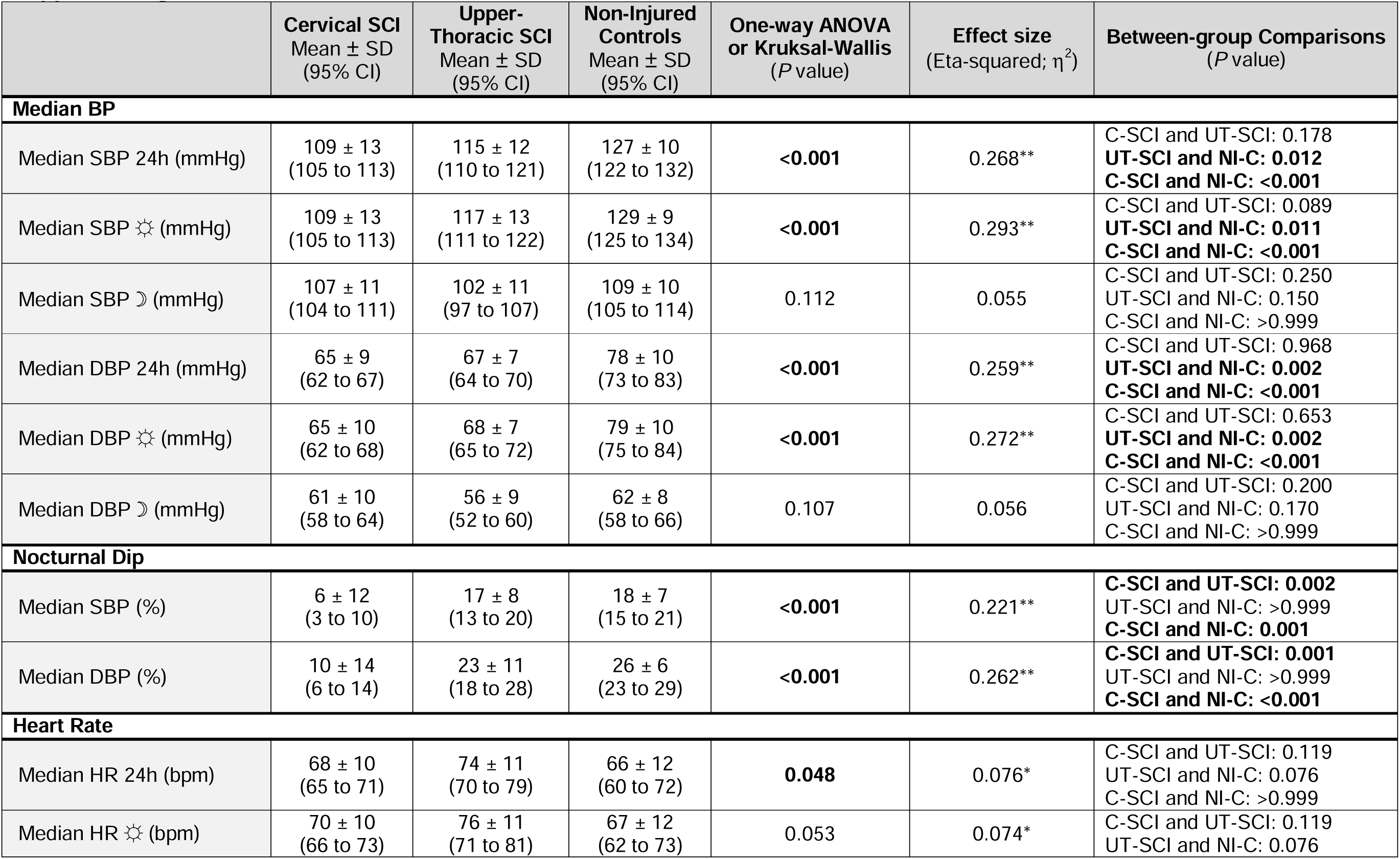
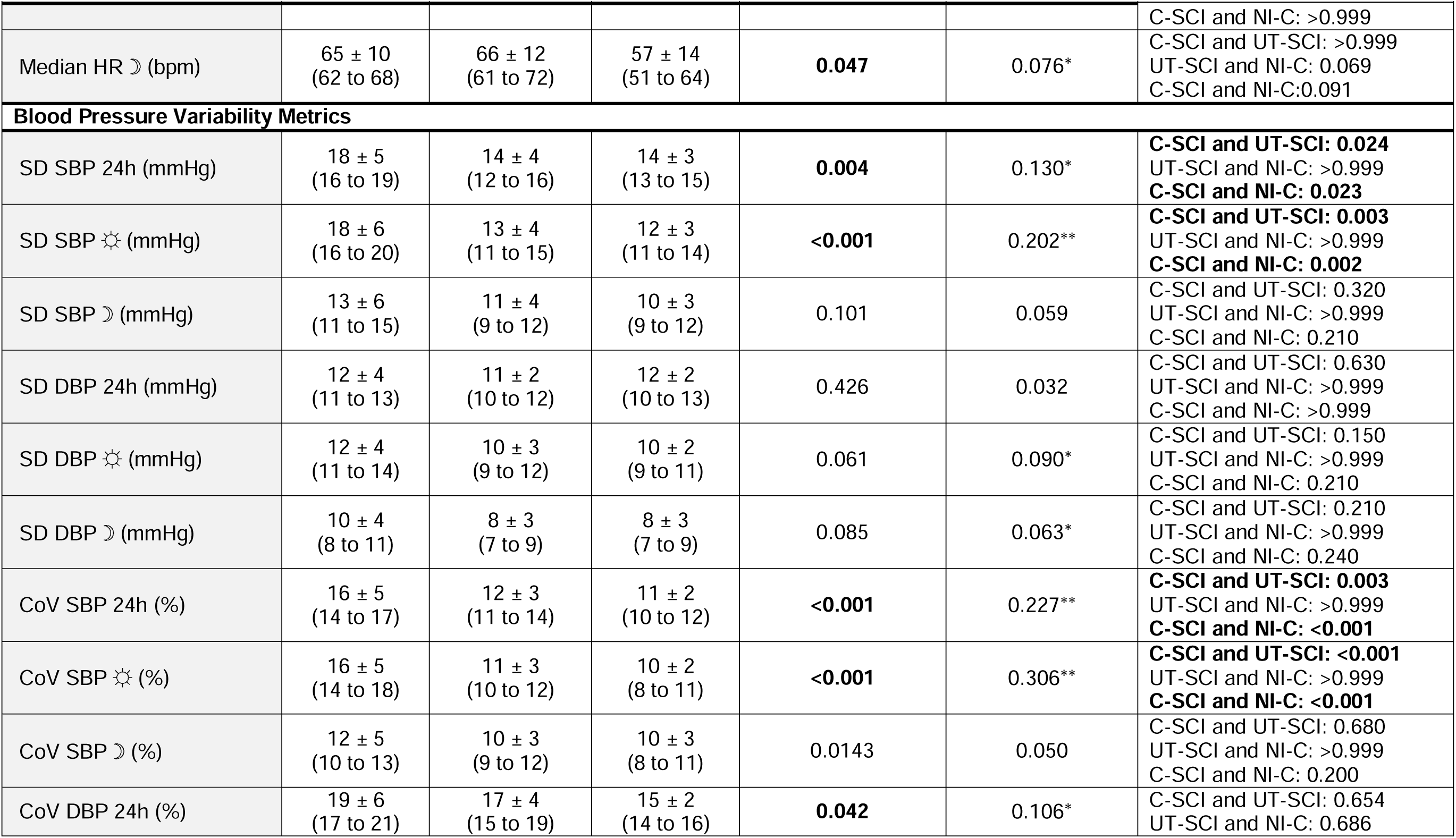
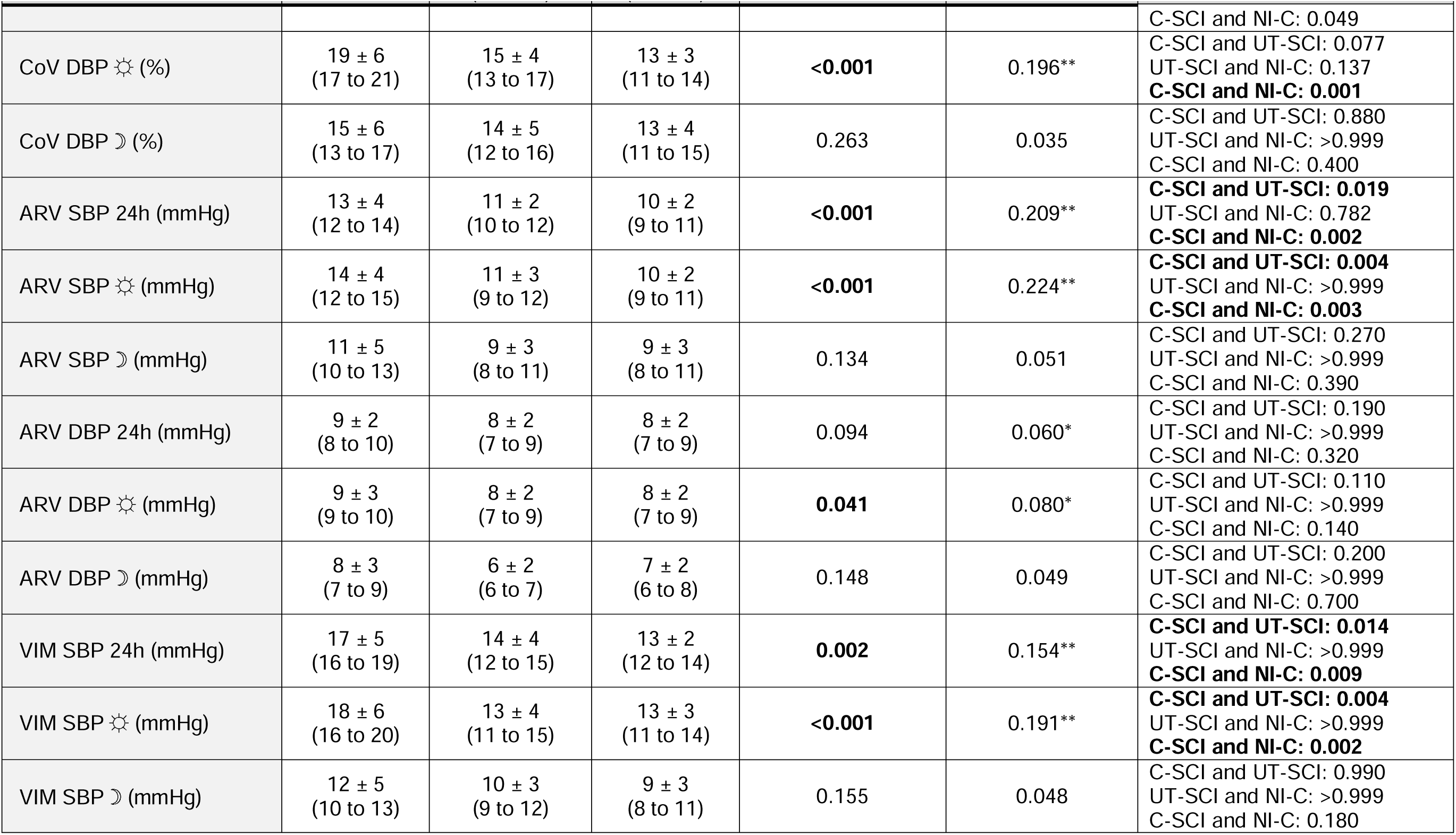
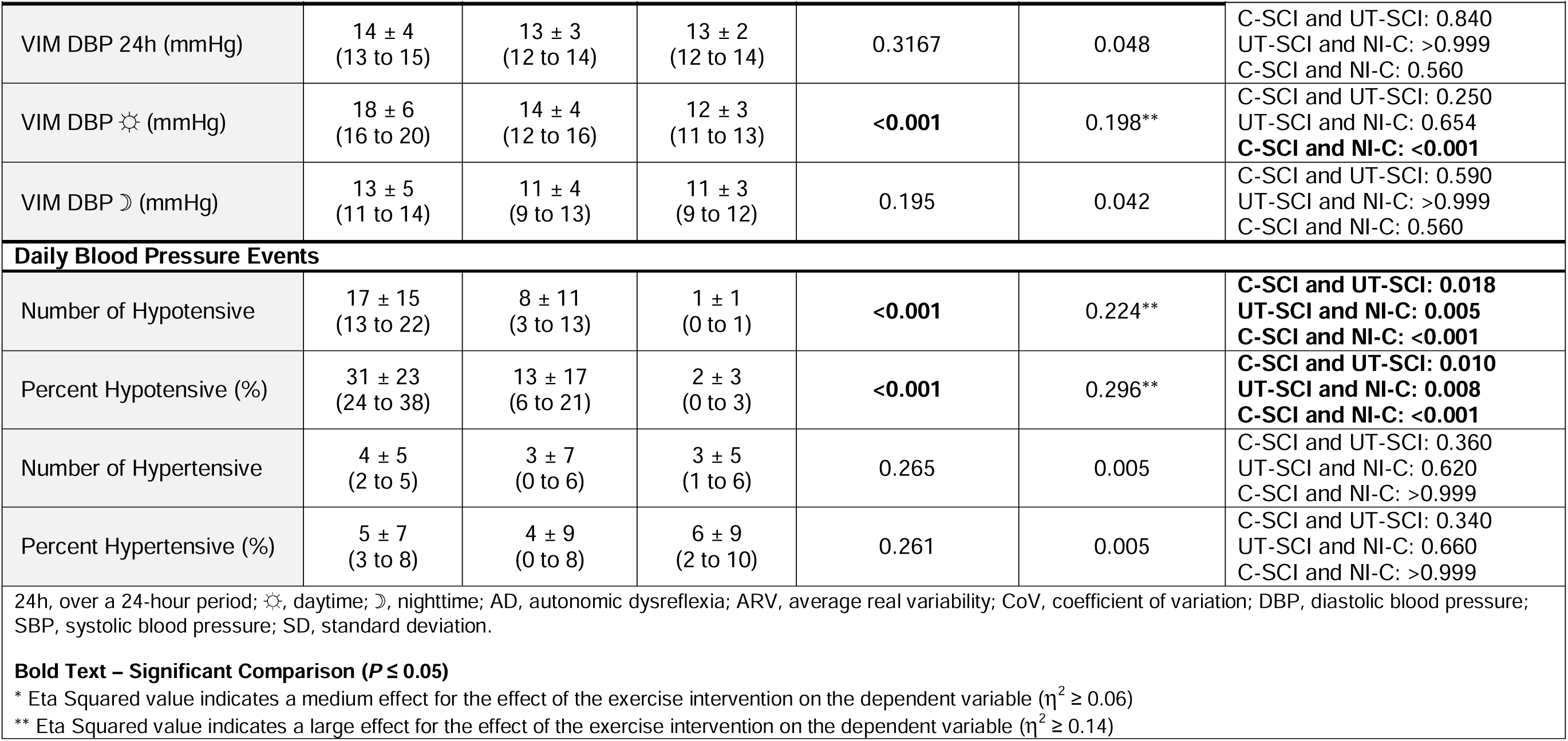

**Supplementary Table 3.** Daytime systolic blood pressure data were resampled to hourly intervals using the median value within each hour to match the lower sampling frequency of nighttime data. This approach ensured comparable data density across day and night periods and allowed direct comparison of blood pressure variability (BPV) metrics between full 24-hour and hourly-thinned datasets.

| BPV Metric | C-SCI | UT-SCI | NI-C | Kruskal-Wallis H | P value | Significant Pairwise Comparisons (Bonferroni adjusted) |
| --- | --- | --- | --- | --- | --- | --- |
| SD (mmHg) | 13.5 ± 5.5 | 9.7 ± 3.7 | 9.2 ± 1.8 | 10.24 | 0.006 | C-SCI > UT-SCI ($P = 0.0437$);<br>C-SCI > NI-C ($P = 0.0229$) |
| CoV (%) | 14.7 ± 6.4 | 10.9 ± 3.3 | 7.6 ± 2.0 | 17.88 | <0.0001 | C-SCI > UT-SCI ($P = 0.00017$);<br>C-SCI > NI-C ($P = 0.0011$) |
| ARV (mmHg) | 12.4 ± 4.0 | 9.0 ± 2.6 | 7.7 ± 2.3 | 13.12 | 0.0014 | C-SCI > UT-SCI ($P = 0.0101$);<br>C-SCI > NI-C ($P = 0.0129$) |
| VIM (mmHg) | 13.9 ± 5.2 | 10.6 ± 3.9 | 9.6 ± 1.7 | 9.41 | 0.0091 | C-SCI > UT-SCI ($P = 0.0497$); |

**Supplementary Figure 1.**
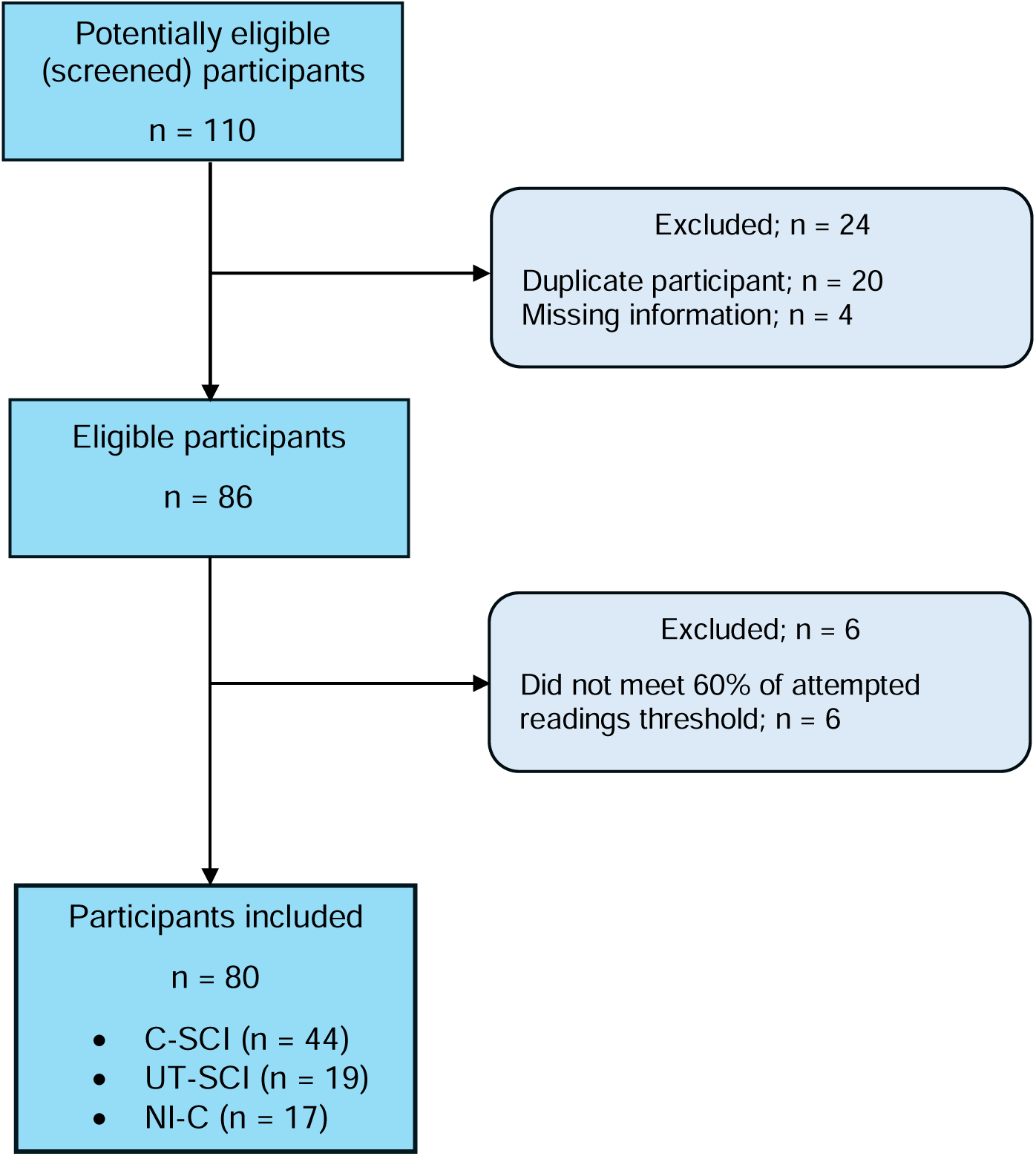
STARD diagram to report flow of participants in the study. The flowchart shows the number of participants at each stage of the study, including those assessed for eligibility, those excluded from the study, and the final number of participants in the analysis. The exclusion criteria were provided with reasons for exclusion to understand the potential sources of bias.

**Supplementary Figure 2**

There was a higher prevalence of non-dippers (SBP and DBP nocturnal dip decrease by 0-10%) and reverse dippers (SBP and DBP nocturnal dip increase) for individuals with cervical spinal cord injury (C-SCI). Dippers (SBP nocturnal dip decrease by 10-20%) were more prevalent in the upper-thoracic spinal cord injury (UT-SCI) and non-injured control (NI-C) groups. Though extreme dippers (SBP and DBP nocturnal dip decrease by ≥20%) were most prevalent in the NI-C group, the young age of the participant cohort would suggest this does not carry a greater cardiovascular risk compared to an older population ^109^.

**Supplementary Figure 2.**
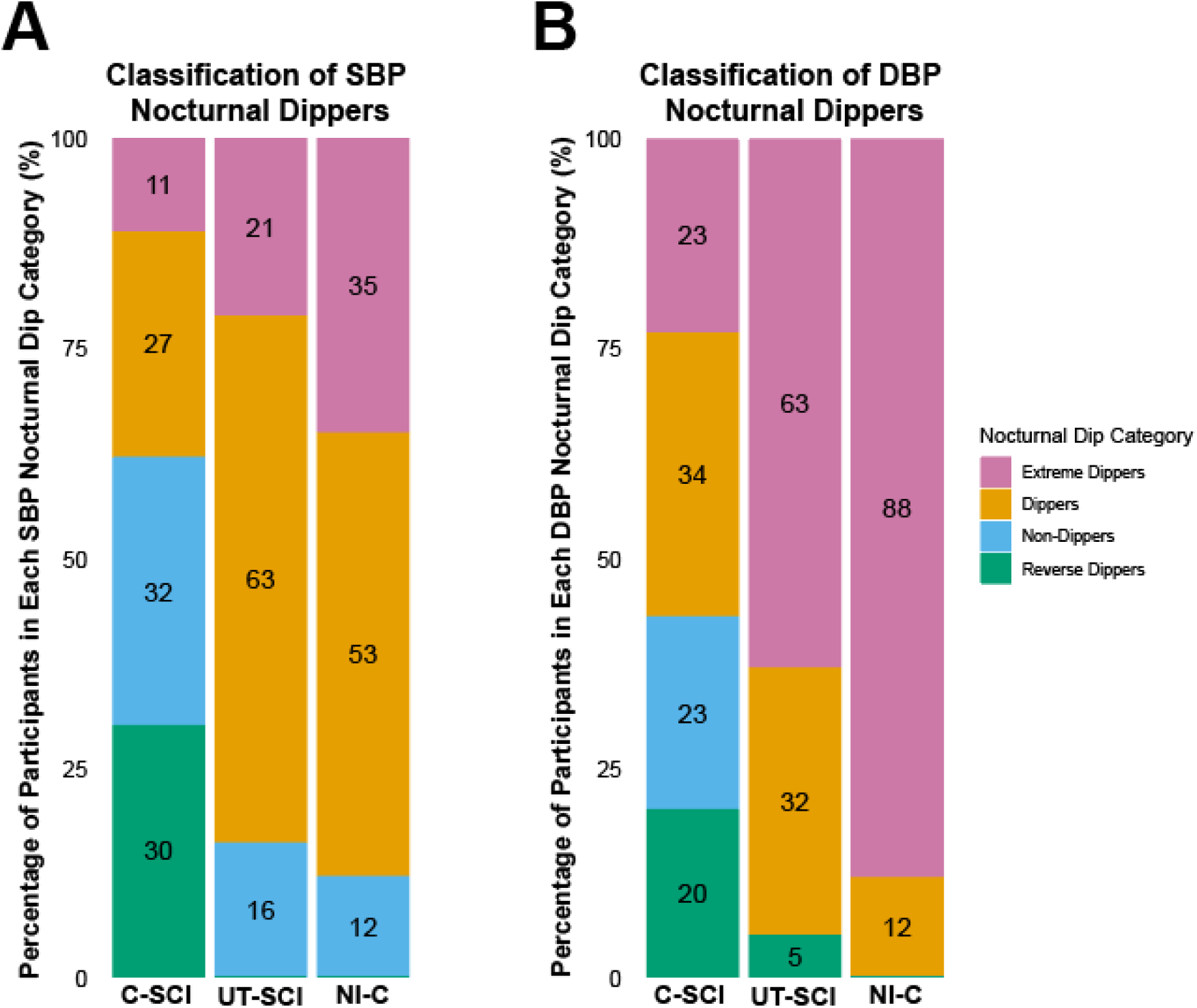
Stacked bar charts of systolic and diastolic nocturnal dip categories. The proportional distribution of individuals within each category of nocturnal dip for each group are displayed. **(A) *Classification of SBP Nocturnal Dippers:*** The prevalence of dippers (orange) was higher in the upper-thoracic spinal cord injury (UT-SCI) and non-injured control (NI-C) groups compared to the cervical spinal cord injury (C-SCI) group, where non-dippers (blue) and reverse dippers (green) were more prevalent. Extreme dippers (purple) were most common in the NI-C group. **(B) *Classification of DBP Nocturnal Dippers:*** The proportion of extreme dippers was highest in the NI-C group, whereas the C-SCI group showed a higher prevalence of reverse dippers and non-dippers. The UT-SCI group had a relatively higher proportion of dippers compared to the C-SCI group. There was a statistically significant association between the DBP nocturnal dip categories and the C-SCI, UT-SCI, and NI-C groups (^2^ = 28.843, *P* < 0.001).

**Supplementary Figure 3**

The URP-CTREE identified homogeneous subgroups based on group (i.e., C-SCI or UT-SCI and NI-C) for all BPV indices (i.e., SD, CoV, ARV, and VIM) of SBP over the 24-hour period (Supplementary Figure 2. A-D). The importance of predictors was assessed using variable importance measures (i.e., how much the variable contributes to the predictive power of the model) for SD (5.50), CoV (0.0014), ARV (3.377), and VIM (3.695), overall indicating that NLI is strongly associated with changes in these BPV indices. Furthermore, NLI as the most important predictor was based on the root mean square error (RMSE; how far predictions are from actual values) and coefficient of determination (*r*^2^; how well the model explains variation in the data) for SD (RMSE = 4.64, *r*^2^ = 0.28), CoV (RMSE = 0.038, *r*^2^ = 0.31), ARV (RMSE = 3.137, *r*^2^ = 0.32), and VIM (RMSE = 4.425, *r*^2^ = 0.26), demonstrating a moderate predictive accuracy.

**Supplementary Figure 3.**
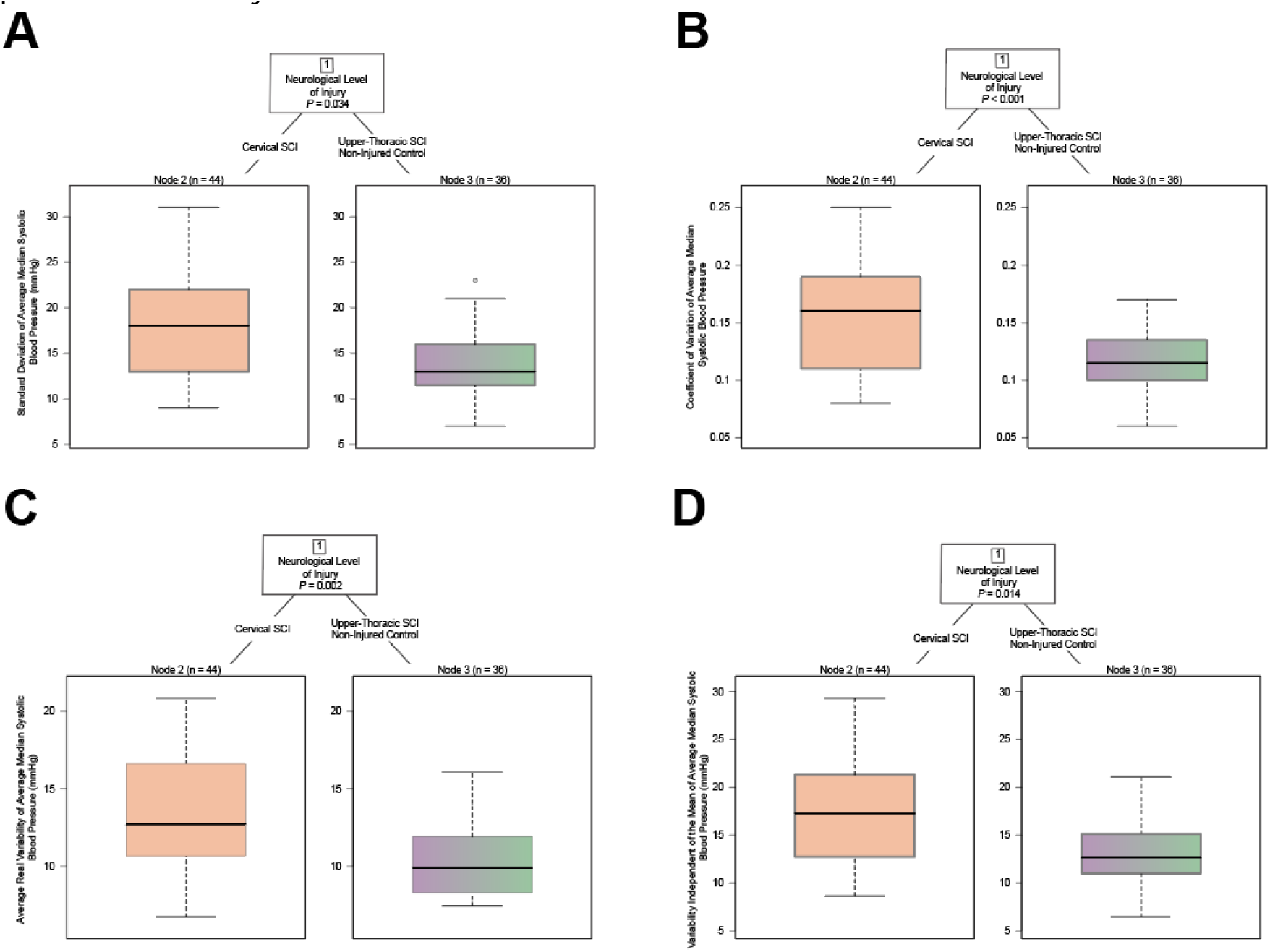
Decision tree models for systolic BPV indices. Conditional inference trees, using a broad set of demographics (i.e., biological sex and age), injury characteristics (i.e., neurological level of injury, injury severity, and time since injury) and study as predictors are displayed over the 24-hour period for **(A)** standard deviation for SBP, **(B)** coefficient of variation for SBP, **(C)** average real variability for SBP, and **(D)** variability independent of the mean for SBP. The upper part represents the sequential splits based on neurological level of injury (cervical spinal cord injury versus upper-thoracic spinal cord injury and non-injured control). Boxplots at the bottom show sample size and distribution of the respective labelled systolic BPV indices within each subgroup (node).

